# Prevalence and Factors Associated with Methicillin-Resistant *Staphylococcus* Nasal Carriage Among People Living with HIV at Kiruddu National Referral Hospital, Kampala, Uganda

**DOI:** 10.64898/2026.05.26.26354086

**Authors:** Juliet Allen Babirye, Freddie Bwanga, Rita Nakalega, Denis Mawanda, Cleopatra Daphne Kugonza, Sharon Miriam Namiiro, Mary Nakiganda, Fred Semitala, Pauline Byakika- Kibwika

## Abstract

Methicillin-resistant *Staphylococcus* (MRS) infections are a significant public health concern. Anterior nares serve as a major reservoir and source of spread of MRS ssp. People living with HIV (PLWHIV) tend to be at higher risk of colonisation with MRS organisms due to frequent healthcare exposure. We assessed the prevalence of MRS nasal carriage and associated factors among PLWHIV at the HIV clinic of Kiruddu National Referral Hospital, Kampala, Uganda, from May to July 2024. Nasal swabs from 256 PLWHIV were cultured, and microbiological isolation was performed at MBN Clinical Laboratories. Prevalence was calculated as proportions, and logistic regression identified associations with clinical and socio-demographic factors (*p* < 0.05). Of 256 participants, 163 (63.7%) carried *Staphylococcus*, with 82 (32%) identified as MRS carriers (8.9% MRSA, 23% MRCoNS). Frequent hospital visits (≥3) (adjusted incidence risk ratio [A-IRR] = 1.18 × 10^7^, *p* < 0.001), second-line antiretroviral therapy (ART) (A-IRR = 3.82, *p* = 0.041), and unsuppressed viral load (>1000 copies/mL) (adjusted odds ratio [AOR] = 11.3, 95% CI: 2.11–60.58, *p* = 0.005) were significantly associated with MRS carriage. Mask-wearing was protective against MRCoNS (A-IRR = 1.66, 95% CI: 1.06–2.58, *p* = 0.026). MRS isolates exhibited high resistance to erythromycin (81.7%) and trimethoprim-sulfamethoxazole (79.3%), but susceptibility to linezolid (93.9%). MRS nasal carriage is prevalent among PLWHIV. Individuals with frequent health care contact and those on second-line ART regimens are more susceptible to MRS colonization, while individuals who wear face masks and those with an undetectable HIV viral load are less susceptible. Antimicrobial Resistance (AMR) surveillance within HIV programs, enhanced infection control, ART adherence, and targeted screening for high-risk groups are critical to mitigate colonization.

## Introduction

Methicillin-resistant *Staphylococcus* (MRS), encompassing *Staphylococcus aureus* (MRSA) and coagulase-negative staphylococci (MRCoNS), are of public health concern due to their ability to cause healthcare-associated and community-acquired infections and to disseminate resistance determinants such as the mecA gene (1-3).

Nasal colonization represents the primary ecological niche for Staphylococcus species and serves as an important reservoir for subsequent infection and transmission in healthcare settings (4). MRS colonization, particularly MRSA, predisposes to infections such as bacteremia, pneumonia, and skin infections, with 20–60% of colonized individuals developing infections (5). MRCoNS, while less pathogenic, can cause severe infections in immunocompromised patients and serve as a reservoir for the *mecA* gene, promoting MRSA emergence (2).

People living with HIV (PLWHIV) are particularly susceptible due to frequent healthcare interactions, repeated antimicrobial exposure (including cotrimoxazole prophylaxis), and underlying immune dysregulation, which may increase the risk of colonization and infection with resistant pathogens and immune suppression (6, 7). Global studies report MRSA nasal carriage rates of 1–16% among PLWHIV, with African data ranging from 2.4–28.2% (8-10).

In Uganda, existing studies have reported relatively low MRSA carriage among PLWHIV with a prevalence of 0–5.2%, and have rarely evaluated MRCoNS colonization, despite its growing relevance in antimicrobial resistance ecology (11, 12). Understanding the burden and determinants of MRS colonization among PLWHIV is important for informing infection prevention strategies, antimicrobial stewardship, and clinical management within HIV care programs. We therefore conducted a cross-sectional study to determine the prevalence of methicillin-resistant *Staphylococcus* nasal carriage and identify associated clinical and socio-demographic factors among PLWHIV attending the HIV clinic at Kiruddu National Referral Hospital in Kampala, Uganda.

## Materials and Methods

### Study Design and Setting

This cross-sectional study was conducted from May to July 2024 at the HIV clinic of Kiruddu National Referral Hospital, a 200-bed tertiary public hospital in Kampala, Uganda, serving approximately 9,600 PLWHIV annually. The nasal swabs were cultured, and microbiological isolation was performed at MBN Clinical Laboratories, a certified facility located 15 km from the hospital, equipped for microbiological testing.

### Study Population, Sampling, and Eligibility Criteria

The target population comprised PLWHIV aged ≥18 years attending the HIV clinic. Using the Kish Leslie formula with a 28.2% MRS prevalence (Muhaba et al., 2022), a sample size of 344 was calculated, adjusted to 256 after accounting for an accessible population of 694 patients and 10% non-response rate. Simple random sampling was employed, with patient chart numbers entered into Excel and randomized using OpenEpi software. Eligible participants were PLWHIV aged ≥18 years who provided written informed consent. Exclusion criteria included recent nasal/facial trauma, surgery, septal/palate defects, or epistaxis.

### Data Collection

The investigator and trained research assistants collected nasal swabs from the anterior nares and clinical/socio-demographic data using pre-tested case report forms (CRFs). Variables included age, sex, education, ART regimen, viral load, and hospital visits. Nasal swabs were collected from both anterior nares using cotton-tipped swabs, placed in Amies transport media (Oxoid, UK), and transported in a triple packaging system to MBN Clinical Laboratories.

### Laboratory Procedures

Swabs were cultured on blood agar, and beta-hemolytic, catalase-positive colonies were subcultured. *S. aureus* was identified by positive tube coagulase and DNase tests, while coagulase-negative staphylococci (CoNS) were catalase-positive but coagulase/DNase-negative. Methicillin resistance was confirmed using cefoxitin disc diffusion (30 μg/mL) on Mueller-Hinton agar, per Clinical and Laboratory Standards Institute (CLSI) guidelines (13). Antibiotic susceptibility was tested for penicillin, erythromycin, clindamycin, gentamicin, tetracycline, chloramphenicol, ciprofloxacin, trimethoprim-sulfamethoxazole, and linezolid.

### Data Analysis

Prevalence was calculated as the proportion of MRS-positive participants. Descriptive statistics used percentages/frequencies for categorical variables and mean (SD) or median (IQR) for continuous variables. Bivariate logistic regression identified factors associated with MRS carriage (*p* < 0.2 for multivariate inclusion). Multivariate logistic regression with stepwise selection calculated adjusted incidence risk ratios (A-IRR) and odds ratios (AOR), with significance at *p* < 0.05. Analyses were conducted using Stata v15.0.

### Ethical Considerations

This study received ethical approval from the School of Medicine Research Ethics Committee (SOMREC) and administrative clearance from Kiruddu Hospital. Written informed consent was obtained, and data were anonymized and stored securely on password-protected devices.

Participants with MRS-positive results were referred to clinicians for management, though decolonization was not indicated per Infectious Diseases Society of America guidelines.

## Results

### Participant Characteristics

Of 256 enrolled PLWHIV, the mean age was 39.9 ± 12.8 years, 67.6% were female, and 53.1% resided in urban areas. Most (51.6%) had primary education, and 48.8% were married. Clinically, 97.7% were on tenofovir-based ART, 65.6% had viral loads <1000 copies/mL, and 93.8% had no recent hospitalizations. Mask-wearing was reported by 61.7%, and 65.2% routinely washed hands at the clinic (Tables 1, 2).

**Table 1:**
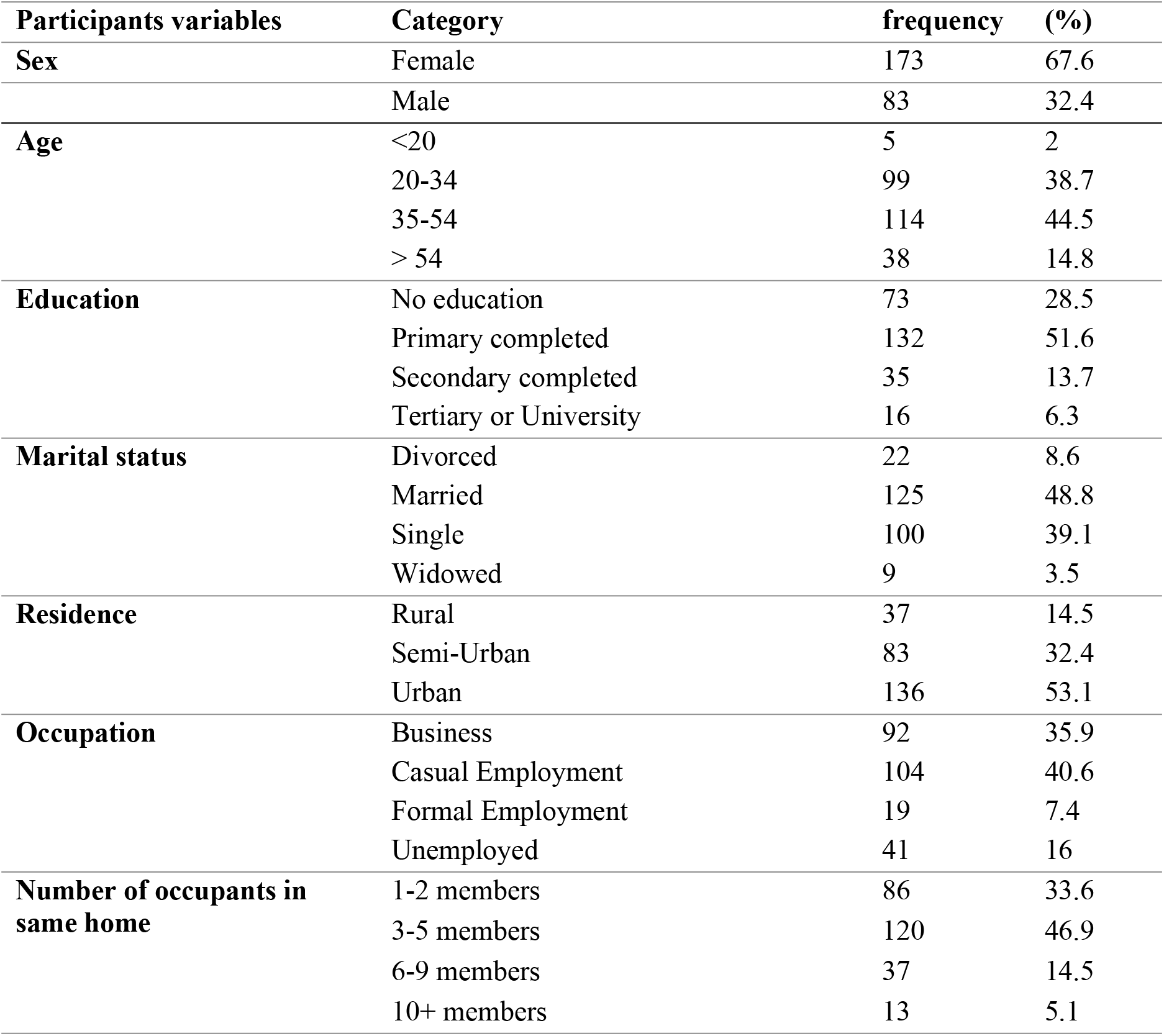
Socio-demographic characteristics of study participants (N=256)

**Table 2:** Clinical and environmental factors (N=256)

| Variable/Category | Frequency (%) |
| --- | --- |
| Currently a cigarette/ tobacco smoker | 10 (3.9) |
| Currently consume alcohol | 55 (21) |
| Currently, inject illicit drugs | 5 (2) |
| Wears a mask in the clinic | 158 (61.7) |
| Awareness of a handwashing facility in the clinic | 194(75.8) |
| Routinely wash hands in the clinic | 167(65.2) |
| Had hospitalized household member (s) in the last 12 months | 10(3.9) |
| Baseline ART (TDF-based regimen) | 242 (94.5) |
| Current ART regimen (TDF/3TC/DTG) | 250(97.7) |
| Had hospital admission in the last 12 months | 16 (6.3) |
| Antibiotic use in the last 6 months | 27 (10.5) |
| Most recent HIV Viral Load <1000 | 168 (65.6) |
| Prior use of Cotrimoxazole prophylaxis | 244(95.3) |

### Prevalence of MRS Nasal Carriage

Bacterial growth occurred in 230 (89.8%) samples, with *Staphylococcus* isolated in 163 (63.7%) participants. Methicillin-resistant *Staphylococcus* nasal carriage was detected in 82 of 256 participants (32%). Among these isolates, 23 (8.9%) were MRSA, and 59 (23%) were MRCoNS.Of the *S. aureus* isolates (n=52), 44.2% were MRSA; of CoNS isolates (n=111), 53.1% were MRCoNS (Table 3).

**Table 3:** Prevalence of bacterial isolates (N=256)

| Isolated Mirco-organisms |  | No. of strains (%) |
| --- | --- | --- |
| Coagulase-negative staphylococcus |  | 111 (43.4) |
| Staphylococcus aureus |  | 52 (20.3) |
| Other organisms |  | 67 (26.1) |
| **Growth** | **Yes** | 230 (89.8) |
|  | **No** | 26 (10.2) |

### Antibiotic Resistance Patterns

MRS isolates showed high resistance to erythromycin (81.7%), trimethoprim-sulfamethoxazole (79.3%), ciprofloxacin (73.2%), and gentamicin (64.6%), but were susceptible to linezolid (93.9%) and chloramphenicol (72%) (Table 4). MRSA and MRCoNS exhibited similar resistance patterns, with MRCoNS showing higher resistance to ciprofloxacin (79.7% vs. 56.5%) and erythromycin (86.4% vs. 69.6%) (Table 5).

**Table 4:** Antibiotic resistance patterns of MRS (n=82)

| Isolated micro-organism | Antibiotic/ Drug |
| --- | --- |
| Staphylococcal species (163) | Cefoxitin |
| Susceptible, n (%) | 77 (30) |
| Resistant, n (%) | 82 (**32**) |
| Intermediate, n (%) |  |
| Missing, n (%) | 4 (1.6) |
| *Staphylococcus aureus* (52) |  |
| Susceptible, n (%) | 29 (11.3) |
| Resistant, n (%) | 23 (**8.9**) |
| Intermediate, n (%) |  |
| Missing, n (%) |  |
| Coagulase-negative Staphylococcus (111) |  |
| Susceptible, n (%) | 48(18.8) |
| Resistant, n (%) | 59(23) |
| Intermediate, n (%) |  |
| Missing, n (%) | 4(1.6) |

**Table 5:**
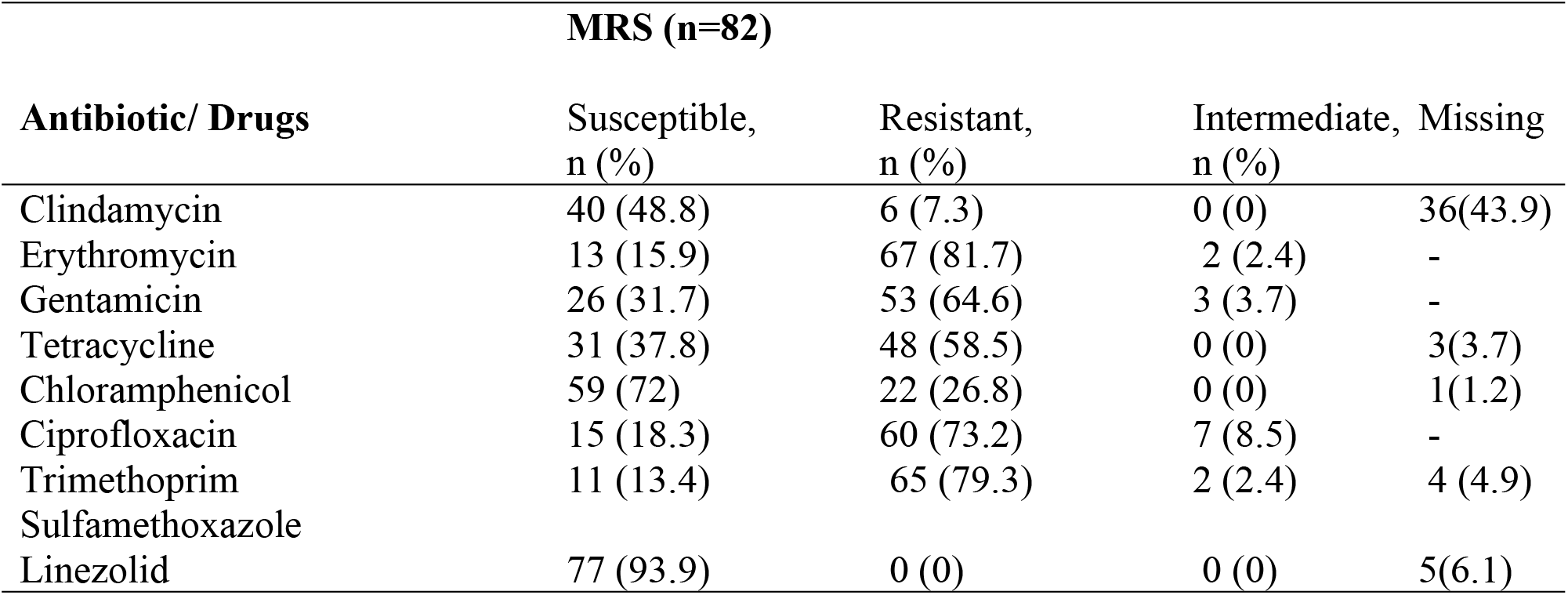
Antibiotic resistance of MRSA vs. MRCoNS)

### Factors Associated with MRS Carriage

Multivariate analysis identified that participants reporting frequent hospital visits (≥3 in 6 months) (A-IRR = 1.18 × 10^7^, *p* < 0.001) and individuals receiving second-line ART (A-IRR = 3.82, *p* = 0.041) had increased risk of MRS colonization. Unsuppressed HIV viral load (>1000 copies/mL) was associated independently with MRSA colonization (AOR = 11.3, 95% CI: 2.11–60.58, *p* = 0.005). Conversely, participants reporting consistent mask use within the clinic environment had lower odds of MRCoNS colonization (A-IRR = 1.66, 95% CI: 1.06– 2.58, *p* = 0.026) (Tables 6–8).

**Table 6:** Multivariate analysis for MRS.

| **Isolated organisms** |  |  |  |  |  |  |  |  |
| --- | --- | --- | --- | --- | --- | --- | --- | --- |
| **Antibiotic/ Drug** | **MRSA(n=23)** |  |  |  | **MR CoNS (n=59)** |  |  |  |
|  | **S,**<br>**n (%)** | **R,**<br>**n (%)** | **I,**<br>**n (%)** | **Missing**<br>**n (%)** | **S,**<br>**n (%)** | **R,**<br>**n (%)** | **I,**<br>**n (%)** | **Missing**<br>**n (%)** |
| **Clindamycin** | 11(47.8) | 2 (8.7) | 0 (0) | 10 (43.5) | 29<br>(49.2) | 4 (6.9) | 0 (0) | 26 (44.1) |
| **Erythromycin** | 7 (30.4) | 16<br>(69.6) | 0 (0) | – | 6 (10.2) | 51<br>(86.4) | 2 (3.4) | – |
| **Gentamicin** | 10 (43.5) | 13<br>(56.5) | 0 (0) | – | 16<br>(27.1) | 40<br>(67.8) | 3 (5.1) | – |
| **Tetracycline** | 13 (56.5) | 10<br>(43.5) | 0 (0) | – | 18<br>(30.5) | 38<br>(64.4) | 0 (0) | 3 (5.1) |
| **Chloramphenicol** | 21 (91.3) | 2 (8.7) | 0 (0) | – | 38<br>(64.4) | 20<br>(33.9) | 0 (0) | 1 (1.7) |
| Ciprofloxacin | 7 (30.4) | 13<br>(56.5) | 3 (13.1) | – | 8 (13.6) | 47<br>(79.7) | 4 (6.8) |  |
| Trimethoprim-sulfamethoxazole | 5 (21.7) | 18<br>(78.3) | 0 (0) | – | 6 (10.2) | 47<br>(79.7) | 2 (3.4) | 4 (6.8) |
| Linezolid | 22 (95.7) | 0 (0) | 0 (0) | 1(4.3) | 55<br>(93.2) | 0 (0) | 0 (0) | 4 (6.8) |

**Table 7:** Multivariate analysis for MRSA.

| Variables | *p-value* | Crude IRR (CI) | *p-value* | Adjusted IRR (CI) |
| --- | --- | --- | --- | --- |
| **No. of hospital visits (within 6 months)** |  |  |  |  |
| Low (1-2 visits) |  | 1 |  | **1** |
| Moderate (3-4 visits)/high (5+ visits) | 0.168 | 1.28(0.90-1.83) | ***p*<0.001** | **1.18e7(2.74e6-5.07e7)** |
| **ART Regimen line** |  |  |  |  |
| First line |  | 1 |  | **1** |
| second line | 0.07 | 0.72(0.50-1.03) | **0.041** | **3.82(1.05-13.81)** |

**Table 8:** Multivariate analysis for MRCoNS.

| Variable & category | *p-value* | COR (CI) | *p-value* | AOR (CI) |
| --- | --- | --- | --- | --- |
| **Body Mass Index (BMI)** |  |  |  |  |
| Normal Weight (18.5-24.49) |  | 1 |  | 1 |
| Underweight (<18.5) | 0.335 | 1.98(0.49-7.96) | 0.464 | 1.75 (0.39-7.96) |
| Overweight (24.5-29.9) | 0.080 | 0.31(0.09-1.15) | 0.096 | 0.31 (0.08-1.23) |
| Obese (≥ 30) | 0.719 | 1.23(0.40-3.73) | 0.938 | 1.05 (0.31-3.61) |
| **Most recent Viral Load** |  |  |  |  |
| <1000 |  | 1 |  | 1 |
| >1000 | **0.005** | **9.48(1.98-45.46) \*** | **0.005** | **11.3 (2.11-60.58) \*** |
| Not done | 0.083 | 2.26(0.90-5.67) | 0.309 | 1.66 (0.63-4.38) |

## Discussion

This study demonstrates that nearly one-third of PLWHIV attending a large urban HIV clinic in Kampala carried methicillin-resistant *Staphylococcus* species in their anterior nares. Although the prevalence of MRSA was relatively modest (8.9%), the overall burden of methicillin-resistant organisms was driven largely by MRCoNS colonization (23%), highlighting the importance of considering coagulase-negative staphylococci within antimicrobial resistance surveillance frameworks.

The observed MRSA prevalence is broadly consistent with reports from other settings in sub-Saharan Africa and Asia, where nasal carriage among PLWHIV has ranged between approximately 2% and 12% (9-10, 12, 14-17). A study conducted in Ghana at a teaching hospital in 2018 reported an MRSA nasal colonization prevalence of 8.2% (9), and in a cross-sectional study in China, the prevalence of nasal carriage of MRSA among PLWHIV was 11.95% (14). Our study’s findings were higher than the 2.4% reported in Ethiopia (15), 3.4% in Taiwan (16), and 5.02% in a tertiary health facility in Buikwe district, Uganda (12), but lower than those reported in Ethiopia at 16.8% and 28% (17, 10). These differences across settings may be influenced by variations in population characteristics, sampling locations, specimen types, and techniques for bacterial isolation and identification.

Our study noted a higher prevalence of MRCoNS colonization. MRCoNS is a source of the mecA gene, which confers methicillin resistance and has the potential to contribute to the emergence of MRSA by transferring the mecA gene from MRCoNS to methicillin-susceptible S. aureus (MSSA) (2). Independently, MRCoNS colonization predisposes to infections such as bloodstream infections, endocarditis, urinary tract infections, surgical site infections, and prosthetic joint infections, particularly in immunocompromised patients, the elderly, critically ill individuals, and those with inserted foreign bodies like catheters (18). Our findings align with a study among 1001 HIV-infected patients in a tertiary facility at Guangzhou, China, which reported 11.89% MRSA and 34.45% MRCoNS prevalence (14). That study also noted multidrug resistance, with MRCoNS isolates showing 81.94% resistance compared to 18.06% for MRSA. Increasing recognition of MRCoNS as both opportunistic pathogens and sources of resistance gene transfer suggests that their epidemiology deserves greater attention in immunocompromised populations.

Frequent healthcare contact (moderate (3–4) or high (≥5) hospital visits within a year of enrollment) emerged as an important predictor of colonization (IRR = 1.18 × 10^7^, p < 0.001). Individuals reporting multiple clinic visits had a substantially higher risk of MRS carriage, consistent with evidence linking repeated healthcare exposure to colonization with antimicrobial-resistant organisms (6,19). HIV programs increasingly utilizing differentiated service delivery models, such as community-based care, could decrease healthcare exposure and MRS colonization among PLWHIV. Strengthening infection prevention practices within outpatient HIV clinics may therefore represent an important component of antimicrobial resistance control.

We also observed that individuals receiving second-line antiretroviral therapy had an increased risk of MRS colonization (IRR = 3.82, p = 0.041). This association may reflect a subgroup of patients with more complex clinical histories, including prolonged healthcare exposure, treatment failure, or greater antibiotic exposure. Similarly, unsuppressed HIV viral load was associated with MRSA colonization (AOR = 11.3; 95% CI: 2.11–60.58, p = 0.005), consistent with findings from a tertiary hospital in Ethiopia linking MRSA colonization to viral loads >1000 copies/mL (10) and another study among children and adolescents also concluded that higher viral loads increased MRSA risk compared to viral loads <1000 copies/mL (20). These findings suggest that immunologic vulnerability may contribute to susceptibility to resistant bacterial colonization.

Interestingly, self-reported mask use in clinic settings was associated with lower MRCoNS colonization. While this finding should be interpreted cautiously due to potential behavioral confounding and self-report bias, it raises the possibility that routine infection prevention practices introduced during the COVID-19 pandemic may have broader benefits for limiting bacterial transmission within healthcare environments.

The high levels of resistance observed to commonly used antibiotics, including erythromycin, ciprofloxacin, and trimethoprim-sulfamethoxazole, are concerning. Our documented cotrimoxazole resistance prevalence of 79.3% was consistent with previous findings from two systematic reviews (21,22). Given the widespread use of cotrimoxazole prophylaxis among PLWHIV in many settings (95.3% of PLWHIV in our study reported prior use), selective pressure for resistant organisms may contribute to this pattern. Continued surveillance of antimicrobial susceptibility patterns in HIV care populations is therefore essential.

This study has several limitations. First, the cross-sectional design precludes causal inference regarding risk factors for colonization. Second, laboratory identification of methicillin resistance relied on phenotypic methods without molecular confirmation. Finally, data were obtained from a single urban referral centre and may not be generalizable to other settings.

Despite these limitations, our findings highlight the substantial burden of methicillin-resistant *Staphylococcus* colonization among PLWHIV in an urban African setting. Integrating antimicrobial resistance surveillance within HIV care programs, strengthening infection prevention measures, and promoting antimicrobial stewardship may help mitigate the risk of colonization and subsequent infection.

## Conclusion

MRS nasal carriage affects one-third of PLWHIV at Kiruddu Hospital. Individuals with frequent health care contact and those on second-line ART regimens are more susceptible to MRS colonization, while those who wear face masks in clinic settings and those with an undetectable HIV viral load are less susceptible. AMR surveillance within HIV programs, enhanced infection control, promoting ART adherence, and targeted screening for high-risk groups are essential to mitigate colonization and prevent infections.

## Data Availability

The datasets generated and analyzed during this study are available from the corresponding author upon reasonable request, subject to ethical restrictions.

## Declarations

### Ethics approval and consent to participate

The study was approved by the School of Medicine Research Ethics Committee (SOMREC), Mak-SOMREC-2023-826. Administrative clearance was received from Kiruddu Hospital.

Written informed consent was obtained from all participants.

### Consent for publication

Not applicable

### Authors’ contributions

JAB, FB, FS, DM, PBK: Conceptualization, methodology, data collection, formal analysis, writing of the original draft.

JAB, FB, FS, PBK: Methodology, data collection, writing, review, and editing.

JAB, FB, FS, PBK: Laboratory analysis, validation, writing, review, and editing.

JAB, FB, RN, DM, CDK, SMN, MN, FS, PBK: Supervision, project administration, writing, review, and editing. All authors read and approved the final manuscript.

## Acknowledgements

We thank the staff at Kiruddu National Referral Hospital and MBN Clinical Laboratories for their support.

## Supplementary Material

The Case Report Form (CRF) is available upon request from the corresponding author.

## References

1) Lowy FD. Staphylococcus aureus infections. New England journal of medicine. 1998 Aug 20;339(8):520–32.

2) Becker K, Heilmann C, Peters G. Coagulase-negative staphylococci. Clinical microbiology reviews. 2014 Oct;27(4):870–926.

3) Haddad SF, Lahr BD, Patarroyo SS, Chesdachai S, Kies KD, O’Horo JC, DeSimone DC, Sendi P, Baddour LM. Bloodstream infection due to coagulase-negative staphylococci: impact of species on prevalence of infective endocarditis. Antibiotics. 2023 Sep 18;12(9):1453.

4) Von Eiff C, Becker K, Machka K, Stammer H, Peters G. Nasal carriage as a source of Staphylococcus aureus bacteremia. New England Journal of Medicine. 2001 Jan 4;344(1):11–6.

5) Simor AE, Loeb M, CIDS/CAMM guidelines committee. The management of infection and colonization due to methicillin-resistant Staphylococcus aureus: a CIDS/CAMM position paper. Canadian Journal of Infectious Diseases and Medical Microbiology. 2004;15(1):39–48.

6) Guaraldi G, Orlando G, Zona S, Menozzi M, Carli F, Garlassi E, Berti A, Rossi E, Roverato A, Palella F. Premature age-related comorbidities among HIV-infected persons compared with the general population. Clinical infectious diseases. 2011 Dec 1;53(11):1120–6.

7) Popovich KJ, Weinstein RA, Aroutcheva A, Rice T, Hota B. Community-associated methicillin-resistant Staphylococcus aureus and HIV: intersecting epidemics. Clinical Infectious Diseases. 2010 Apr 1;50(7):979–87.

8) Sabbagh P, Riahi SM, Gamble HR, Rostami A. The global and regional prevalence, burden, and risk factors for methicillin-resistant Staphylococcus aureus colonization in HIV-infected people: A systematic review and meta-analysis. American journal of infection control. 2019 Mar 1;47(3):323–33.

9) Boison D, Akwetey SA, Osei SA, Kelechi S, Barnie PA. Nasal colonization of methicillin-resistant Staphylococcus aureus in HIV-infected patients at the Cape Coast Teaching Hospital, Ghana. Frontiers in Tropical Diseases. 2022 Sep 20;3:976567.

10) Muhaba H, Fenta GM, Gebretsadik D. Methicillin resistance Staphylococcus aureus nasal carriage and its associated factors among HIV patients attending art clinic at Dessie comprehensive specialized hospital, Dessie, North East Ethiopia. PLOS Global Public Health. 2022 Sep 12;2(9):e0000838.

11) Bebell LM, Ayebare A, Boum Y, Siedner MJ, Bazira J, Schiff SJ, Metlay JP, Bangsberg DR, Ttendo S, Firth PG. Prevalence and correlates of MRSA and MSSA nasal carriage at a Ugandan regional referral hospital. Journal of Antimicrobial Chemotherapy. 2017 Mar 1;72(3):888–92.

12) Ssenyonga B, Mwebaze S, Atuhairwe C, MugishaTaremwa I. Perspective for Methicillin-resistant Staphylococcus aureus colonization, antibiotic susceptibility patterns and risk factors for colonization among people living with HIV at Nyenga hospital, Buikwe district, in central Uganda. International Journal of Infection Prevention. 2018 Jul 31;1(1):1–8.

13) Clinical and Laboratory Standards Institute CL. Performance standards for antimicrobial susceptibility testing.

14) Li Y, Lin J, Li L, Cai W, Ye J, He S, Zhang W, Liu N, Gong Z, Ye X, Yao Z. Methicillin-Resistant Coagulase-Negative Staphylococci Carriage is a Protective Factor of Methicillin-Resistant Staphylococcus Aureus Nasal Colonization in HIV-Infected Patients: A Cross-Sectional Study. Canadian Journal of Infectious Diseases and Medical Microbiology. 2021;2021(1):5717413.

15) Gebremedhn G, Gebremariam TT, Wasihun AG, Dejene TA, Saravanan M. Prevalence and risk factors of methicillin-resistant Staphylococcus aureus colonization among HIV patients in Mekelle, Northern Ethiopia. Springerplus. 2016 Jun 24;5(1):877.

16) Hsu YY, Wu D, Hung CC, Huang SS, Yuan FH, Lee MH, Huang CT, Shie SS, Huang PY, Yang CC, Cheng CW. Methicillin-resistant Staphylococcus aureus nasal colonization among HIV-infected patients in Taiwan: prevalence, molecular characteristics and associated factors with nasal carriage. BMC infectious diseases. 2020 Mar 30;20(1):254.

17) Lemma MT, Zenebe Y, Tulu B, Mekonnen D, Mekonnen Z. Methicillin resistant Staphylococcus aureus among HIV infected pediatric patients in Northwest Ethiopia: carriage rates and antibiotic co-resistance profiles. PLoS One. 2015 Sep 30;10(9):e0137254.

18) Michalik M, Samet A, Podbielska-Kubera A, Savini V, Miedzobrodzki J, Kosecka-Strojek M. Coagulase-negative staphylococci (CoNS) as a significant etiological factor of laryngological infections: a review. Annals of clinical microbiology and antimicrobials. 2020 Jun 4;19(1):26.

19) Maciel RA, Klück HM, Durand M, Sprinz E. Comorbidity is more common and occurs earlier in persons living with HIV than in HIV-uninfected matched controls, aged 50 years and older: a cross-sectional study. International Journal of Infectious Diseases. 2018 May 1;70:30–5.

20) Siberry GK, Frederick T, Emmanuel P, Paul ME, Bohannon B, Wheeling T, Barton T, Rathore MH, Dominguez KL. Methicillin-Resistant Staphylococcus aureus Infections in Human Immunodeficiency Virus-Infected Children and Adolescents. AIDS Research and Treatment. 2012;2012(1):627974.

21) Olaru ID, Tacconelli E, Yeung S, Ferrand RA, Stabler RA, Hopkins H, Aiken AM, Kranzer K. The association between antimicrobial resistance and HIV infection: a systematic review and meta-analysis. Clinical Microbiology and Infection. 2021 Jun 1;27(6):846–53.

22) Assefa M, Girmay G. Prevalence of co-trimoxazole resistance among HIV-infected individuals in Ethiopia: a systematic review and meta-analysis. Frontiers in Medicine. 2024 Jul 11;11:1418954.

